# Extending influenza surveillance to detect non-influenza respiratory viruses of public health relevance: analysis of surveillance data, 2015-2019, Belgium

**DOI:** 10.1101/2021.01.13.20202200

**Authors:** Lorenzo Subissi, Nathalie Bossuyt, Marijke Reynders, Michèle Gérard, Nicolas Dauby, Patrick Lacor, Siel Daelemans, Bénédicte Lissoir, Xavier Holemans, Koen Magerman, Door Jouck, Marc Bourgeois, Bénédicte Delaere, Sophie Quoilin, Steven Van Gucht, Isabelle Thomas, Cyril Barbezange

**Affiliations:** National Influenza Centre, Sciensano, Brussels, Belgium; European Public Health Microbiology Training Programme (EUPHEM), European Centre for Disease Prevention and Control, Stockholm, Sweden; Epidemiology of Infectious Diseases, Sciensano, Brussels, Belgium; Department of Laboratory Medicine, Medical Microbiology, Algemeen Ziekenhuis Sint-Jan, Brugge-Oostende AV, Belgium; Centre Hospitalier Universitaire St-Pierre, Brussels, Belgium; Centre for Environmental Health and Occupational Health, School of Public Health, Université Libre de Bruxelles (ULB), Brussels, Belgium; Internal Medicine-Infectious Diseases, Universitair Ziekenhuis Brussel, Brussels, Belgium; Pediatric Pulmonary and Infectious Diseases, Universitair Ziekenhuis Brussel, Brussels, Belgium; Microbiology, Grand Hôpital de Charleroi, Charleroi, Belgium; Infectiology, Grand Hôpital de Charleroi, Charleroi, Belgium; Clinical Laboratory, Jessa Ziekenhuis, Hasselt, Belgium; Infection Control, Jessa Ziekenhuis, Hasselt, Belgium; Centre Hospitalier Universitaire UCL Namur, Ysoir, Belgium

**Author notes:** Corresponding author: Cyril Barbezange, Corresponding author.

**Keywords:** severe acute respiratory infection, influenza-like illness, human metapneumovirus, coronavirus, respiratory syncytial virus

## Abstract

**BACKGROUND:** Seasonal influenza-like illness (ILI) affects millions of people yearly. Severe acute respiratory infections (SARI), mainly caused by influenza, are a leading cause of hospitalisation and mortality. Increasing evidence indicates that non-influenza respiratory viruses (NIRVs) also contribute to the burden of SARI. In Belgium, SARI surveillance by a network of sentinel hospitals is ongoing since 2011.

**AIM:** Here, we report the results of using in-house multiplex PCRs for the detection of a flexible panel of viruses in respiratory ILI and SARI samples and the estimated incidence rates of SARI associated to each virus.

**METHODS:** ILI was defined as an infection with onset of fever and cough or dyspnoea. SARI was defined as an infection requiring hospitalization with onset of fever and cough or dyspnoea within the previous 10 days. Samples were collected during four winter seasons and tested by multiplex RT-qPCRs for influenza virus and NIRVs. Using catchment population estimates, incidence rates of SARI associated to each virus were calculated.

**RESULTS:** One third of the SARI cases were positive for NIRVs, reaching 49.4% among children under fifteen. In children under five, incidence rates of NIRV-associated SARI were double that of influenza (103.4 versus 57.6 per 100000 person-months), with NIRV co-infections, respiratory syncytial viruses, human metapneumoviruses and picornaviruses contributing the most (33.1, 13.6, 15.8 and 18.2 per 100000 person-months, respectively).

**CONCLUSION:** Early testing for NIRVs could be beneficial to clinical management of SARI patients, especially in children under five, for whom the burden of NIRV-associated disease exceeds that of influenza.

## INTRODUCTION

Acute viral infections of the respiratory tract are common in humans. According to the World Health Organization (WHO), complications such as lower respiratory tract infections and pneumonia are among the main causes of mortality in children and the elderly worldwide [1]. The burden attributed to seasonal influenza virus has long received most of the attention [2], but the involvement of non-influenza respiratory viruses (NIRVs), such as respiratory syncytial virus and human metapneumovirus, is increasingly documented [3,4]. However, the burden of NIRVs [5–7] compared to seasonal influenza [8] is rarely estimated. These data would allow to better understand the need for enhanced surveillance of these viruses during winter seasons with the aim of improving patient management. In December 2019, a new NIRV (Severe Acute Respiratory Syndrome Coronavirus 2, or SARS-CoV2) emerged in Wuhan (China) causing a pandemic, and as of June 3^rd^ 2020 has infected over 6.1 million people and killed over 370000 people across the vast majority of countries and territories of the world.

The influenza surveillance network was implemented in 1952 by WHO to monitor the continuous antigenic changes of the virus and to guide vaccine composition. Following the 2009 pandemic, the scope of the surveillance was extended to better assess the severity and burden of influenza viruses. In Belgium, influenza virus surveillance is organised by the national public health institute, Sciensano, which also hosts the National Influenza Centre (NIC). Two sentinel networks are in place: i) surveillance of influenza-like illness (ILI) in the community, based on general practitioners providing information on mild cases (in place since the mid-1970s); ii) surveillance of severe acute respiratory infection (SARI) based on six sentinel hospitals providing information on hospitalised patients (set up after the 2009 influenza pandemic and operating every season since 2011). Since the 2015/2016 influenza season, the NIC introduced the systematic testing of NIRVs by RT-qPCRs in addition to testing and characterizing influenza viruses. We report here the results of systematic NIRV testing of ILI and SARI surveillance samples for four seasons (2015/2016 to 2018/2019) and we estimate the burden and severity of NIRVs, as compared to influenza.

## METHODS

### Settings, Participants and Variables

Sentinel-based ILI surveillance was performed through a network of general practitioners (GPs) widely distributed throughout Belgium and representing approximately 1.5% of all Belgian GPs. Patients were enrolled during each influenza season surveillance period (from week 40 to week 20). Recommendations were that nasopharyngeal swabs were obtained weekly from the first two ILI patients belonging to two different households. SARI patients were recruited through the sentinel network of six hospitals in Belgium, two in each administrative region of the country: Flanders, Wallonia and Brussels-Capital. Paediatric and adult units were recommended to systematically collect both detailed clinico-epidemiological data and a respiratory specimen from all patients meeting the SARI case definition during the epidemic period of seasonal influenza (starting and ending weeks between January and April, depending on influenza activity). Based on WHO’s guidelines, ILI and SARI cases were defined as acute respiratory illness with onset within the last 10 days, with measured or reported fever of ≥38°C and with cough and/or dyspnoea. In addition, overnight hospitalisation was required for SARI cases. Patients for whom informed consent was not obtained (either directly or from a parent or legal guardian) were excluded from the study. Standardized questionnaires were used to collect data on age, sex, clinical signs included in the case definition, status of vaccination against influenza viruses, administration of a neuraminidase inhibitor antiviral and/or antibiotic treatment, and comorbidities known as risk factors of severity. Follow-up data during hospitalisation were also reported for the SARI cases to evaluate disease severity and included the detection of pneumonia based on chest radiography, and/or the development of acute respiratory distress syndrome (ARDS), and/or the requirement for respiratory assistance and/or for extracorporeal membrane oxygenation (ECMO), and/or the admission in intensive care unit (ICU), and/or death (all-cause death).

### Laboratory testing

Respiratory samples were analysed at Belgian National Influenza Centre. Viral nucleic acids were extracted using BioMerieux’s NucliSENS EasyMag. The following respiratory virus targets were detected using several in-house multiplex RT-qPCRs: influenza viruses types A and B (and subsequent subtype/lineage), respiratory syncytial viruses types A and B, human metapneumoviruses, parainfluenzaviruses types 1, 2, 3 and 4, coronaviruses CoV-OC43, CoV-NL63 and CoV-229E, adenoviruses, picornaviruses (*rhinovirus* and *enterovirus* genera), specific enterovirus D68, parechovirus, and bocavirus (adapted from original protocols by US CDC; LJ van Elden, University Medical Centre Utrecht, The Netherlands; P Overduin, RIVM, The Netherlands; O Hungnes and K Bragstad, Institute of Public Health, Norway). Primer/probe sequences and PCR conditions are available upon request.

### Burden of disease of virus-associated SARI

To estimate the burden of disease of virus-associated SARI, the catchment population estimates from 2017 (mid-year of the study period) were used to calculate monthly incidence rate per 100,000 population during the winter season, based on WHO recommendations [9]. Catchment populations of the SARI hospitals by age group were estimated based on the hospital admission data (i.e. the proportion of the different municipalities) and the total population of these municipalities; this rough estimation has the advantage to be easily extracted without extra working load for the hospitals. The number of months covered by the study period (i.e. 14.9) used for the denominator were calculated by adding up the number of weeks with active SARI surveillance for each season (i.e. 64), multiplied by 7 and divided by the average number of days in a month during the winter period (December-April, i.e. 30). Age group-specific catchment populations of the six hospitals were summed up to calculate the monthly incidence rate by age group (<5, 5-14, 15-64 and ≥65 years old) for virus-associated SARI. 95% Confidence intervals were calculated using the Rothman-Greenland method [10].

### Ethics approval

The SARI surveillance protocol was approved by a central Ethical Committee (reference AK/12-02-11/4111; in 2011: Centre Hospitalier Universitaire St-Pierre, Brussels, Belgium; from 2014 onwards: Universitair Ziekenhuis Brussel, Brussels, Belgium) and the local ethical committees of each participating hospital. Informed consent was obtained from all participants or parents/guardians.

## RESULTS

### Study population

Over the four seasons under study, 1791 ILI and 4774 SARI patients responding to the case definition were retained in the analysis (Figure 1). Among the ILI patients with known age, 78.7% were adults (15-64 years old, 1356/1724), 14.1% were children (<15 years old, 244/1724) and 7.2% were older adults (65 years old and above, 124/1724). Half were females (833/1692), including 11 who were pregnant. Among the SARI patients, there were slightly more males than females (52.2%; 2428/4650), with 28 females being pregnant. Children and older adults represented 35.5 % and 44.1% of SARI cases with known age, respectively. Among children, 44.6% and 42.0% were less than 1 year old (753/1689) and between 1 and 4 years old (711/1689), respectively. The six sentinel hospitals contributed heterogeneously to the distribution of SARI patients in terms of age, with one hospital reporting more paediatric cases (Supplement Table S1).

**Figure 1:**
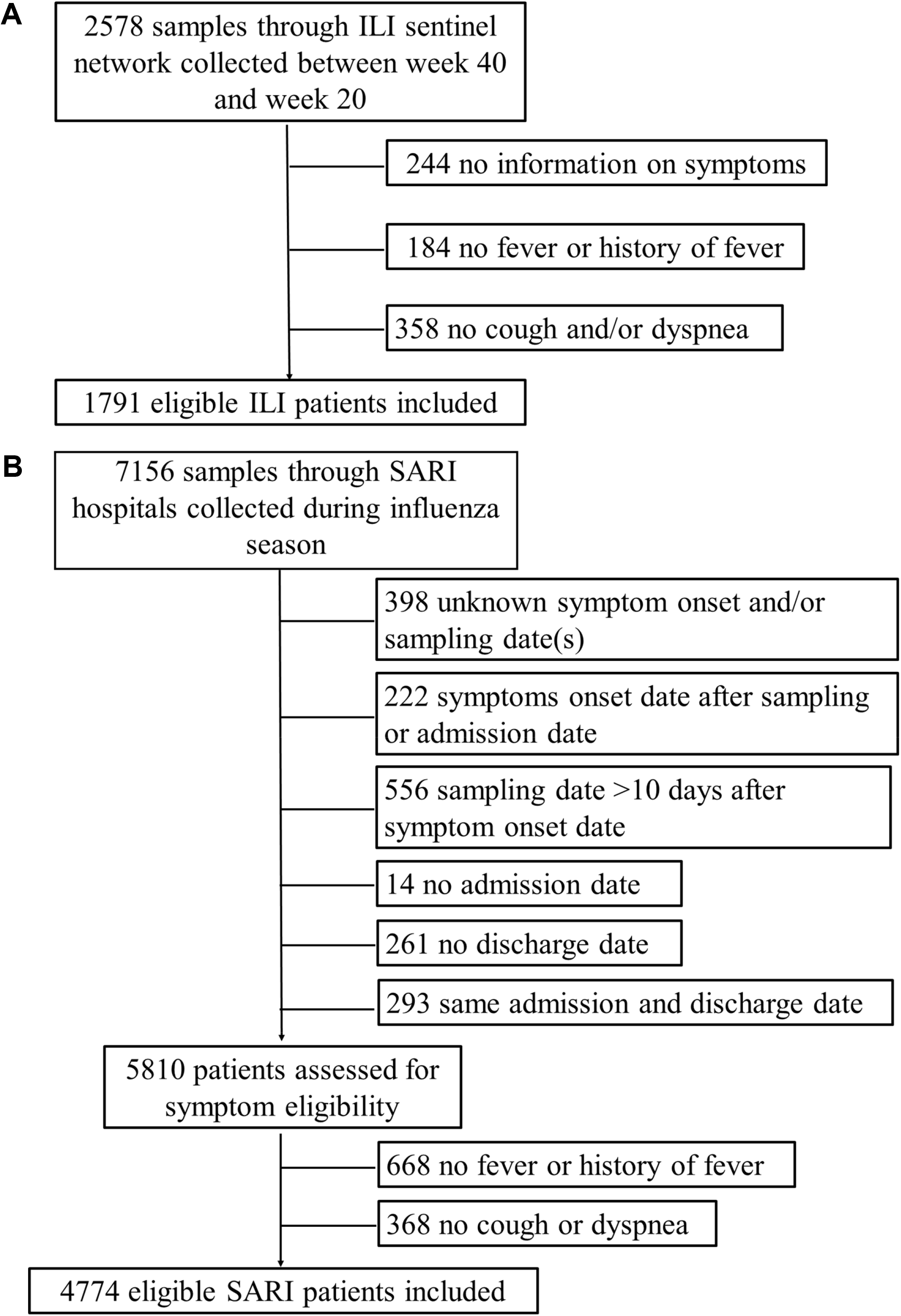
Flow diagram for participant inclusion in the study. (A) for influenza-like illness (ILI) surveillance; (B) for severe acute respiratory infection (SARI) surveillance.

### Overall respiratory virus detection rates

Among the 1791 ILI patients, 408 were negative for all respiratory viruses tested, 1068 were positive for influenza virus (of which 81 were co-detections with non-influenza respiratory viruses, NIRVs), and 293 were negative for influenza virus but positive for a NIRV (Figure 2A). Among the 4774 SARI patients, 1309 were negative for all respiratory viruses tested, 2049 were positive for influenza virus (of which 250 were co-detections with NIRVs) and 1125 were negative for influenza virus but positive for at least one NIRV (Figure 2B).

**Figure 2:**
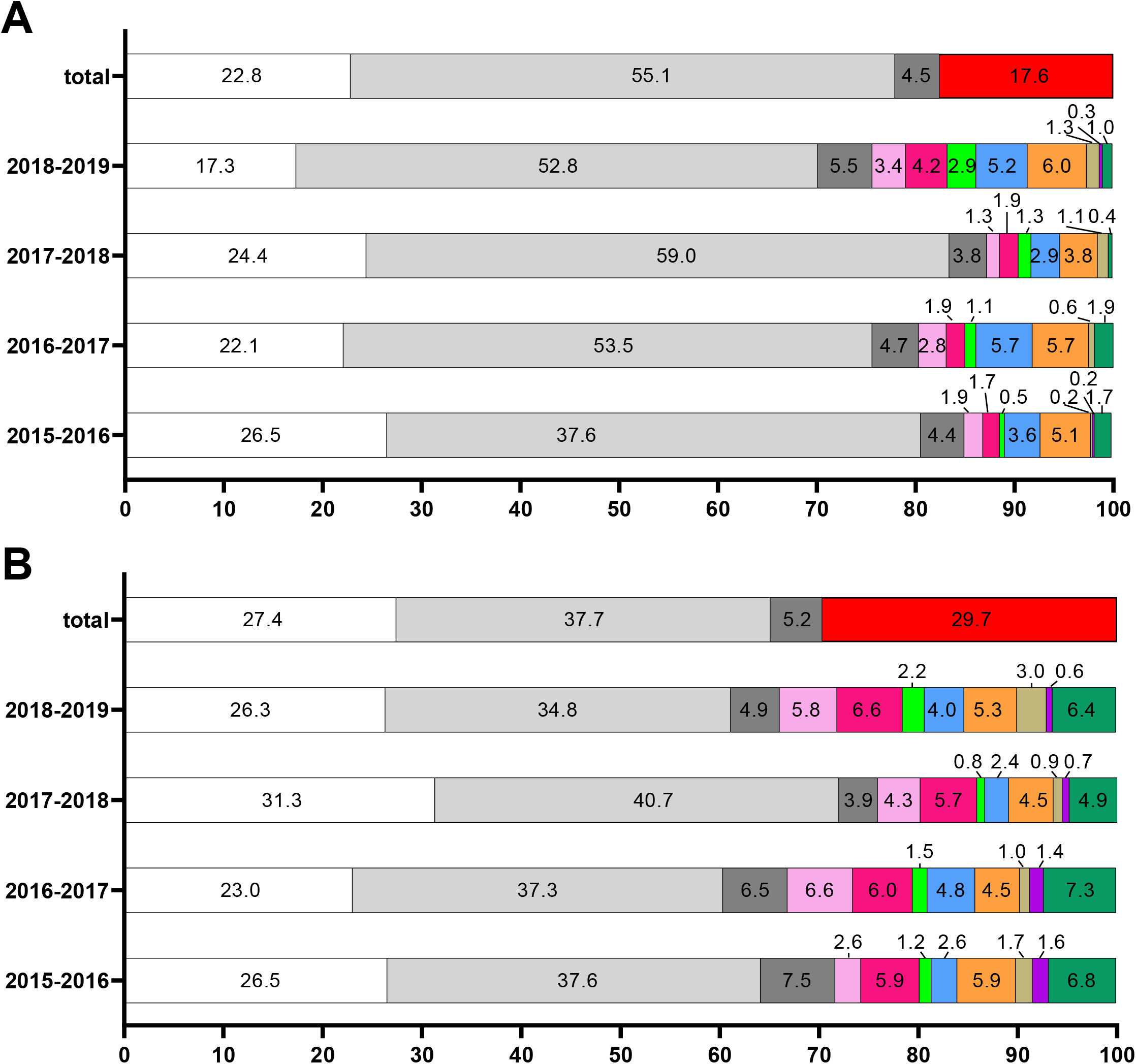
Comparison of the proportions of the different respiratory virus detected by multiplex RT-qPCRs per season and overall. Colour codes: white, negative; light grey, influenza virus type A or B; dark grey, co-detection of influenza virus with at least one non-influenza respiratory virus; red, positive for non-influenza respiratory virus(es); light pink, Respiratory Syncytial virus type A or B; fuchsia, human metapneumovirus; green, parainfluenzavirus type 1, 2, 3 or 4; blue, coronavirus CoV-OC43, CoV-NL63 or CoV-229E; orange, picornavirus of the *rhinovirus* and *enterovirus* genera or parechovirus; light brown, adenoviruses; purple, bocavirus; dark green, co-detection of several non-influenza respiratory viruses. (A) for influenza-like illness (ILI) surveillance; (B) for severe acute respiratory infection (SARI) surveillance.

The most common single NIRV detections among ILI patients were picornaviruses (5.1%, 91/1791) and coronaviruses (4.3%, 77/1791), followed by hMPV (2.3%, 42/1791) and RSV (2.3%, 41/1791). Parainfluenza virus, adenovirus and bocavirus single detections accounted for 1.4%, 0.8%, and 0.1%, respectively. Co-detections of at least two viruses accounted for 5.7% of the ILI samples (102/1791), with 81 co-detections of influenza virus with NIRVs and 22 co-detections of NIRVs. Among SARI patients, the main single NIRV detections were hMPV (291/4774), RSV (238/4774), picornaviruses (237/4774) and coronaviruses (162/4774) (Figure 2B). Parainfluenza virus, adenovirus and bocavirus single detections accounted for 1.4%, 1.7%, and 0.9%, respectively. Co-detections were more common among SARI patients as compared to ILI patients. They accounted for 11.3% of the overall number of tested SARI samples (541/4,774), with influenza virus-positive (n=250) and influenza-negative (n=291) co-detections representing 5.2 and 6.1%, respectively.

The proportion of NIRV detection (excluding when in association with influenza viruses) varied between 12.8% in 2016-2017 and 24.4% in 2018-2019 in the ILI samples (Figure 2A). In SARI patients, the proportion was higher and varied between 24.2% in 2016-2017 and 33.9% in 2018-2019 (Figure 2B). Within a season, the proportions of picornaviruses and coronaviruses were relatively similar among ILI and SARI patients. On the contrary, RSV and hMPV detection rates were systematically higher in SARI than in ILI samples. Except for season 2015-2016, these two viruses represented the largest groups in SARI patients, whereas picornaviruses and coronaviruses were the most frequent ones in the ILI patients. Season 2018/2019 was marked by higher detection rates for parainfluenza viruses (both in ILI and SARI) and adenoviruses (in SARI), compared to previous seasons (Figure 2).

Both ILI and SARI surveillances were able to capture the seasonal circulation of influenza virus, but such bell-shape epidemic curves were not detected for other NIRVs, with perhaps two exceptions: coronaviruses in the ILI surveillance during season 2016/2017 despite the limited number of positive samples (Supplement Figure S1); and RSV in the SARI surveillance, with the detection of the end of the RSV epidemic, which usually occurs before the influenza epidemic in Western Europe [11] (Figure 3).

**Figure 3:**
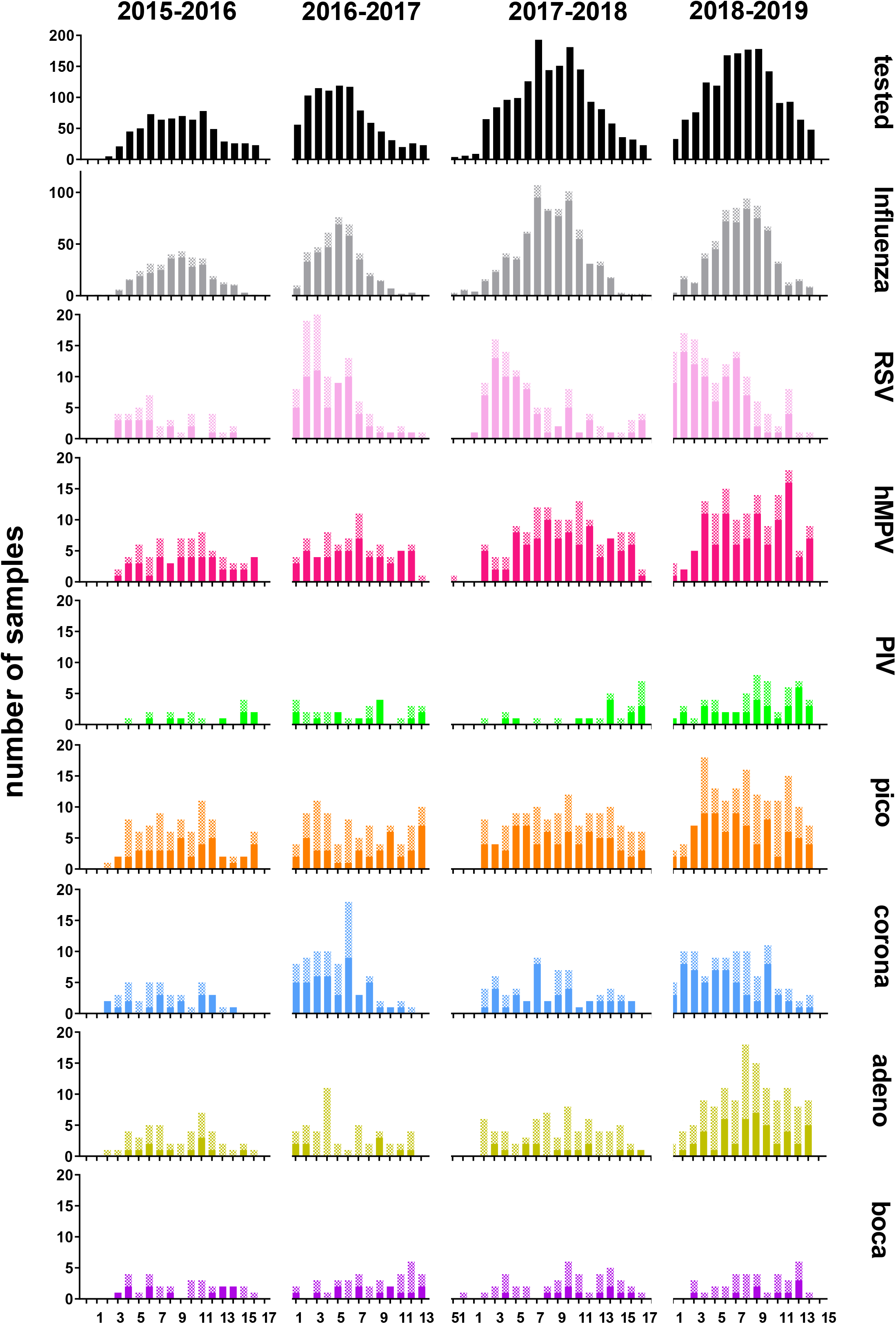
Distribution of the positive samples for each respiratory virus by season and sampling week for SARI patients. Colour codes: black, tested by multiplex RT-qPCRs (Tested); light grey, influenza virus type A or B (Influenza); light pink, Respiratory Syncytial virus type A or B (RSV); fuchsia, human metapneumovirus (hMPV); green, parainfluenzavirus type 1, 2, 3 or 4 (PIV); orange, picornavirus of the *rhinovirus* and *enterovirus* genera or parechovirus (Pico); blue, coronavirus CoV-OC43, CoV-NL63 or CoV-229E (Corona); light brown, adenovirus (Adeno); purple, bocavirus (Boca). Plain bar, single detection; chequered pattern, in co-detection with at least one other virus. X-axis, weeks of active SARI surveillance: 2015-2016, week 1 to week 17 of 2016; 2016-2017, week 1 to week 13 of 2017; 2017-2018, week 51 of 2017 to week 17 of 2018; 2018-2019, week 1 to week 15 of 2019.

### Age-specific NIRV detection rates and NIRV-associated SARI incidence rates

In children (less than 15 years old), the proportion of samples positive for NIRVs (single or co-detection) was higher in SARI than ILI patients. The proportion of the SARI children positive for NIRVs only (excluding co-detection with influenza viruses) was 49.4% (829/1678), and only 15.6% (38/244) of ILI children (Fisher’s exact test, p<0.001). On the contrary, the proportions of positive NIRV samples among adults (15-64 years old) and older adults (≥65 years old) taken together were similar for ILI and SARI surveillance (17.9%, 267/1489, and 18.9%, 582/3084, respectively; Fisher’s exact test, p=0.465).

Half (82/162) of the coronavirus-only positive SARI samples were from older adults (Table 1). On the contrary, all bocavirus-only (n=44) and 75.9% (63/83) of the adenovirus-only positive SARI samples were from children. RSV-only and hMPV-only positive SARI samples were from both children (44.1%, 105/238, and 44.7%, 130/291, respectively), mainly less than 1 year old, and from older adults (42.0%, 100/238, and 38.5%, 112/291, respectively). Picornavirus-only and parainfluenzavirus-only SARI samples were mainly from children less than 5 years old (58.2%, 138/237, and 50.7%, 35/69, respectively), but also from older adults (21.9%, 52/237, and 29.0%, 20/69, respectively). Finally, SARI samples with co-detection of NIRVs or of NIRVs with influenza viruses were largely from children less than 5 years old (86.3%, 251/291, and 51.6%, 129/250, respectively).

**Table 1:**
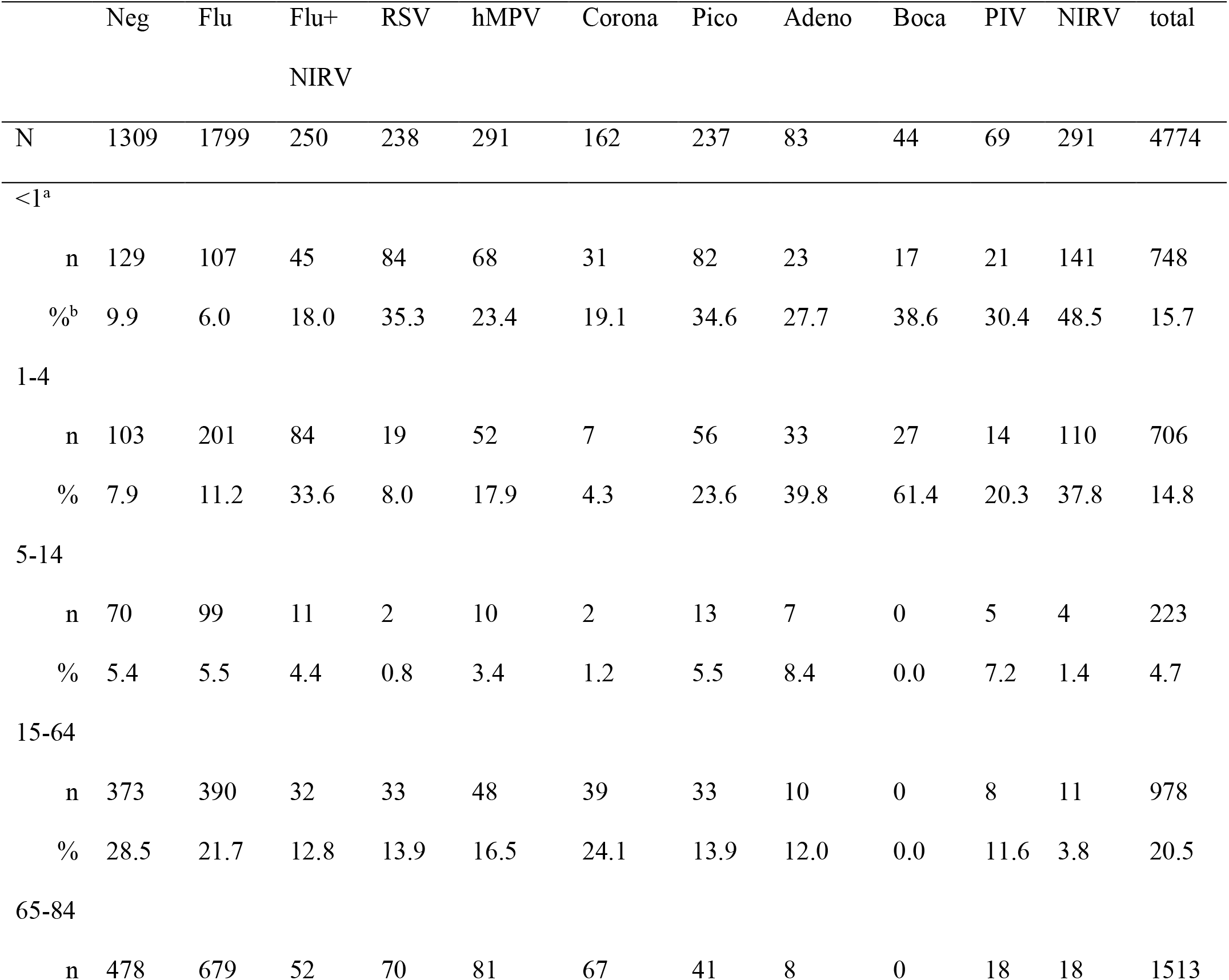
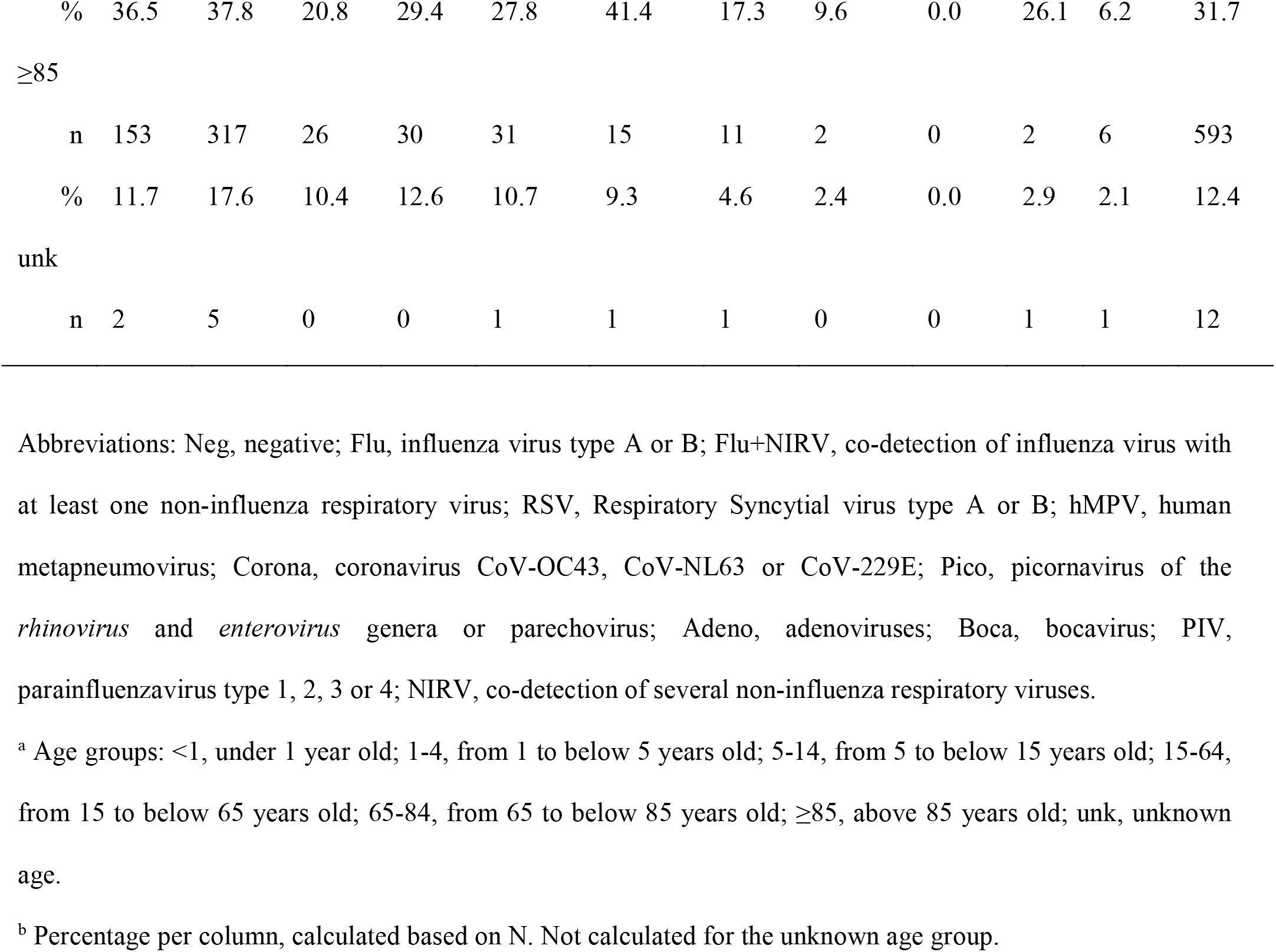
Respiratory virus single and co-detection in SARI patients by age group

The burden of NIRVs captured by influenza surveillance was further evaluated by calculating the incidence rates of SARI associated to each virus within specific age groups and overall (Figure 4). NIRV incidence rates were noticeably higher in the youngest age group (<5 years old) than in the other age groups or the overall population, with RSV, hMPV, and picornaviruses contributing the most as single infection (13.6, 15.8 and 18.2 per 100000 person-months, respectively), followed by adenoviruses, bocavirus, coronaviruses and parainfluenzaviruses (7.4, 5.8, 5.0 and 4.6 per 100000 person-months, respectively). In this age group, the incidence rate of SARI associated to all kind of influenza-negative NIRV infections was almost double that associated to influenza (103.5 versus 57.6 per 100000 person-months). Incidence rates of SARI associated to NIRVs were low in the other age groups, as compared to those associated to influenza (Figure 4). Despite that, the incidence rates of SARI associated to RSV, hMPV and coronaviruses in older adults were nonetheless more than twice that in the overall population (3.9, 4.4, 3.2 versus 1.8, 2.1, 1.2 per 100000 person-months, respectively).

**Figure 4:**
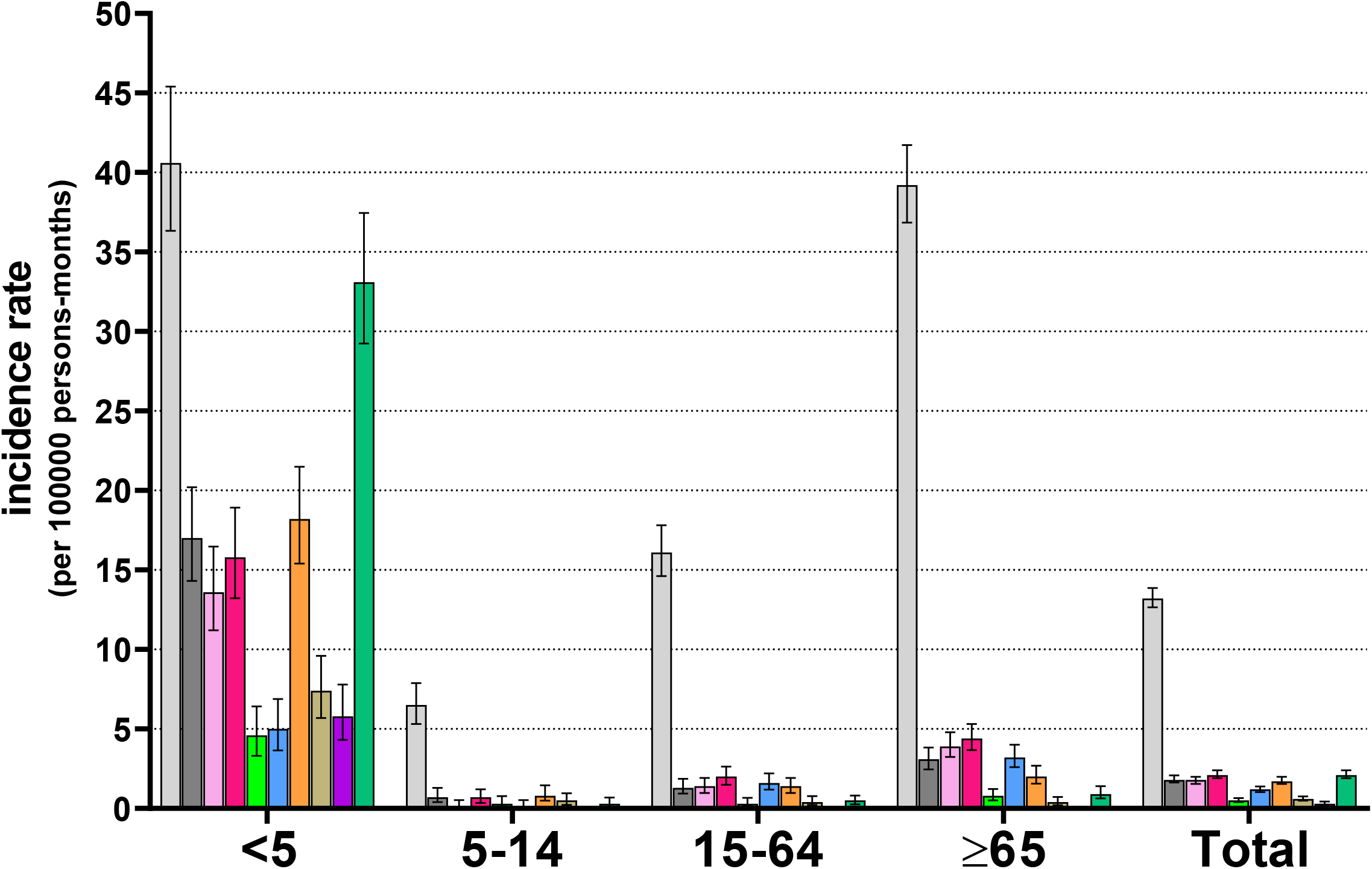
Incidence rates of virus-associated SARI per 100000 person-months by age group and overall. Colour codes: light grey, influenza virus type A or B; dark grey, co-detection of influenza virus with at least one non-influenza respiratory virus; light pink, Respiratory Syncytial virus type A or B; fuchsia, human metapneumovirus; blue, coronavirus CoV-OC43, CoV-NL63 or CoV-229E; orange, picornavirus of the *rhinovirus* and *enterovirus* genera or parechovirus; green, parainfluenzavirus type 1, 2, 3 or 4; purple, bocavirus; light brown, adenoviruses; dark green, co-detection of several non-influenza respiratory viruses. X-axis, age groups: <5, below 5 years old; 5-14, from 5 to below 15 years old; 15-64, from 15 to below 65 years old; ≥65, above 65 years old; Total, whole population.

## DISCUSSION

Here, we report on the use of several in-house multiplex RT-qPCRs on respiratory specimens collected through two networks of influenza surveillance (ILI and SARI) to detect a broad range of respiratory viruses of public health relevance during the winter season. We found that NIRV testing may explain up to one third of infections and becomes extremely useful for diagnostic purposes of paediatric SARI patients. By estimating incidence rates, we showed that NIRVs even represent a higher burden than influenza viruses in the SARI patients under 5 years old.

In Belgium, systematic testing of NIRVs on samples collected through the ILI and SARI surveillance was introduced from the winter season 2015-2016 onwards. The use of in-house multiplex PCRs offers the flexibility to change the panel of viruses/virus groups depending on the situation. With the emergence of SARS-CoV-2, it could be possible to substitute one of the current target with primers and probes for this new coronavirus, using for example the protocols already designed [12], since ILI and SARI surveillance networks will be important tools to follow the SARS-CoV2 epidemics in the coming years.

The first limitation of our study is that these surveillance networks collect data only during the winter season in Belgium. This is particularly true for the SARI network which operates only during the period of influenza virus activity. Therefore, the detection rates observed here for the different NIRVs might not totally represent their whole circulation patterns. For example, it was shown that the intensive circulation period for RSV usually precedes that of influenza viruses in Western Europe [11]. Though RSV, hMPV, coronaviruses and other NIRVs do not strictly follow the same pattern as influenza viruses [13], many tend to also circulate during the winter season in temperate regions, thus making influenza surveillance a relevant system for their study [14,15].

Another limitation is that paediatric and geriatric samples are underrepresented in ILI patients, probably due to sampling bias (GPs in Belgium being less likely to have children or older people as patients or to sample them). The systematic testing for NIRVs allowed the detection of a pathogen in only one eighth of the ILI patients, suggesting that the additional effort of testing NIRVs in this study population might be of limited interest. On the contrary, NIRV positive samples were almost a third of the SARI patients, where the morbidity and mortality associated to respiratory infection is the highest, and almost half in paediatric SARI patients. The burden associated to NIRVs in SARI patients was age-dependent, and particularly high for young children (<5 years old). While NIRV co-infections were rarely detected among ILI patients, they were almost as common as influenza virus infection among paediatric SARI patients under five years old. This clearly shows the benefit that testing SARI patients for NIRV infections can have on the surveillance and clinical management.

RSV and hMPV were common among paediatric SARI patients, especially those younger than five years old. Interestingly, older adults represented another large proportion of RSV infections. The non-negligible burden associated with RSV is important to take into consideration in the coming years, when RSV vaccines will become available and vaccination is implemented. It may be of interest to consider the possibility to add older adults among the high risk groups for which vaccination could be recommended [16]. Preliminary research even suggested that a RSV vaccine could also provide protection against hMPV through the induction of cross neutralizing antibodies [17,18]. This highlights the relevance of the surveillance of both RSV and hMPV in the years following RSV vaccine introduction.

The objective of our SARI surveillance was to evaluate the severity of influenza viruses, and the definition used might not be the most adapted one for NIRVs: the criteria ‘fever’ and ‘cough’ were indeed chosen in an attempt to specifically target influenza viruses [19], however they are also characteristic symptoms of most NIRVs. Interestingly, cough and dyspnoea are clinical signs that were previously found to be positively associated with RSV and hMPV [20], the two main NIRVs identified in our study. Despite the differences in the case definition and the period of surveillance, our results remain comparable to those obtained in other countries regarding the NIRVs identified and their proportions [21,22]. Interactions between respiratory viruses have been reported. For example, a negative association between rhinovirus and RSV infection was reported in children [23]. Similarly, an increased risk of non-influenza virus infections was reported in children who received inactivated influenza vaccine [24]. Through SARI surveillance, it would also be possible to estimate the potential impact of newly circulating viruses such SARS-CoV-2 on the incidence of NIRVs.

Despite an important decrease in the number of specimens that were reported as negative in SARI patients following the introduction of NIRV testing, over one fourth of SARI patients remained negative for all tested viruses. Although viruses are considered as the main cause of acute respiratory tract infections [25–27] and our methodology covers the main respiratory viruses that are involved [28], it is possible that some samples were positive for viruses that were not targeted by our multiplex PCRs (e.g. coronavirus HKU, influenza C virus, cytomegalovirus, herpes simplex virus), or for bacterial or fungal respiratory pathogens, known to substantially contribute to complications in lower respiratory tract infections [29–31]. In a prospective study in 11 European countries that looked at the aetiology of lower respiratory tract infections among adults, a bacterial pathogen was detected in one fifth of the participants [32]. The difference in the proportion of ‘negative’ samples that we observed between age groups (one third among adults and one fifth among children) translates into a different range of pathogens causing SARI in children (more broadly covered by our multiplex PCRs) as compared to adults. An explanation may be that the clinical signs of the SARI case definition might be less specific for adults, especially older adults [33,34].

In conclusion, our study demonstrated the added value of testing NIRVs with a flexible method and confirmed that the burden of disease associated to NIRVs in SARI patients should not be underestimated, especially in children under the age of five, for whom it is even higher than that of influenza viruses.

## Data Availability

Aggregated data are provided in the manuscript and the supplementary materials. For any other data, specific ethical approval might be required and the authors should be contacted.

## Funding

This work was supported by the Belgian Federal Public Service ‘Health, Food Chain Safety, and Environment’, the Belgian National Insurance Health Care (INAMI/RIZIV), and the Regional Health Authorities of Flanders (Agentschap Zorg en Gezondheid, AZG), of Brussels (Commission communautaire commune de Bruxelles-Capitale, COCOM), and of Wallonia (Agence pour une Vie de Qualité, AVIQ).

## Conflict of interest

The authors declare no conflict of interest.

## Acknowledgements

The authors would like to thank Jeannine Weyckmans, Ilham Fdillate, Assia Hamouda and Reinout Van Eycken from the NIC for their excellent technical assistance throughout the surveillance seasons; Evelyn Petit from AZ Sint-Jan, Françoise Antoine, Tiphaine Mouquet and Christelle Nguepi from CHU St-Pierre, Lore-Lien Roels and Prof. Elke De Wachter from UZ Brussels, Marlies Blommen, Annemie Forier and Dr. Luc Waumans from Jessa Ziekenhuis, Catherine Sion and Mohamed El Kaissi from Grand Hôpital de Charleroi for their dedication in the follow-up of the SARI cases; and all the doctors and nurses of the six hospitals who took part in the recruitment of the SARI cases, without whom the SARI surveillance would not be possible. N.D. is a Post-doctorate Clinical Master Specialist of the FRS-FNRS (Fond National de la Recherche Scientifique).

## Contributions

**Project Administration:** C.B., N.B., I.T.; **Data Curation:** L.S., C.B., N.B., I.T.; **Formal Analysis:** L.S., C.B.; **Investigation:** M.R., M.G., N.D., P.L., S.D., B.L., X.H., K.M., D.J., M.B., B.D., C.B., I.T.; **Resources:** M.R., M.G., N.D., P.L., S.D., B.L., X.H., K.M., D.J., M.B., B.D.; **Visualization:** L.S., C.B.; **Writing – Original Draft Preparation:** L.S., C.B.; **Writing – Review & Editing:** all authors; **Funding Acquisition:** S.Q., S.V.G.

## Notes

### Competing Interest Statement

The authors have declared no competing interest.

