## Supplement Table S1 for "Extending influenza surveillance to detect non-influenza respiratory viruses of public health relevance: analysis of surveillance data, 2015-2019, Belgium"

**Supplement Table S1: Characteristics of ILI and SARI patients for influenza seasons 2015-2016, 2016-2017, 2017-2018 and 2018-2019, Belgium.**

| | ILI | | SARI per hospital and total | | | | | | | | | | | | | |
|---|---|---|---|---|---|---|---|---|---|---|---|---|---|---|---|---|
| | | | Site 1 | | Site 2 | | Site 3 | | Site 4 | | Site 5 | | Site 6 | | total | |
| | n | %[b] | n | % | n | % | n | % | n | % | n | % | n | % | n | % |
| Overall | 1791 | | 1071 | | 514 | | 491 | | 832 | | 1034 | | 832 | | 4774 | |
| Age group[a] | | | | | | | | | | | | | | | | |
| <1 | 6 | 0.3 | 107 | 10.0 | 246 | 47.9 | 44 | 8.9 | 110 | 13.2 | 86 | 8.3 | 160 | 19.2 | 753 | 15.8 |
| 1-4 | 41 | 2.3 | 99 | 9.2 | 135 | 26.3 | 40 | 8.2 | 84 | 10.1 | 190 | 18.4 | 163 | 19.6 | 711 | 14.9 |
| 5-14 | 197 | 11.0 | 27 | 2.5 | 28 | 5.5 | 5 | 1.0 | 35 | 4.2 | 67 | 6.5 | 63 | 7.6 | 225 | 4.7 |
| 15-64 | 1365 | 76.2 | 245 | 22.9 | 61 | 11.9 | 141 | 28.7 | 204 | 24.5 | 165 | 16.0 | 158 | 19.0 | 974 | 20.4 |
| 65-84 | 112 | 6.3 | 425 | 39.7 | 33 | 6.4 | 211 | 43.0 | 288 | 34.5 | 344 | 33.3 | 208 | 25.0 | 1509 | 31.6 |
| ≥85 | 12 | 0.7 | 166 | 15.5 | 11 | 2.1 | 47 | 9.6 | 111 | 13.3 | 182 | 17.6 | 73 | 8.8 | 590 | 12.4 |
| Missing | 57 | - | 2 | - | 0 | - | 3 | - | 0 | - | 0 | - | 7 | - | 12 | - |
| Sex | | | | | | | | | | | | | | | | |
| Female | 833 | 49.2 | 526 | 48.3 | 238 | 46.8 | 176 | 44.1 | 391 | 47.5 | 529 | 51.4 | 372 | 45.6 | 2222 | 47.8 |
| Male | 859 | 50.8 | 552 | 51.7 | 271 | 53.2 | 223 | 55.9 | 433 | 52.5 | 501 | 48.6 | 448 | 54.6 | 2428 | 52.2 |
| Missing | 99 | - | 3 | - | 5 | - | 92 | - | 8 | - | 4 | - | 12 | - | 124 | - |
| Season | | | | | | | | | | | | | | | | |
| 2015-16 | 411 | 22.9 | 94 | 8.8 | 119 | 23.2 | 73 | 14.9 | 81 | 9.7 | 190 | 18.4 | 134 | 16.1 | 691 | 14.5 |
| 2016-17 | 474 | 26.5 | 255 | 23.8 | 99 | 19.3 | 96 | 19.6 | 83 | 10.0 | 223 | 21.6 | 156 | 18.8 | 912 | 19.1 |
| 2017-18 | 525 | 29.3 | 363 | 33.9 | 177 | 34.3 | 156 | 31.8 | 311 | 37.4 | 317 | 30.7 | 287 | 34.5 | 1611 | 33.8 |
| 2018-19 | 281 | 15.7 | 359 | 33.5 | 119 | 23.2 | 166 | 33.8 | 357 | 42.9 | 304 | 29.4 | 255 | 30.7 | 1560 | 32.7 |

[a] Age groups: <1, under 1 year old; 1-4, from 1 to below 5 years old; 5-14, from 5 to below 15 years old; 15-64, from 15 to below 65 years old; 65-84, from 65 to below 85 years old; ≥85, above 85 years old; unk, unknown age.

[b] Percentage per column, calculated based on overall. Not calculated for 'missing' groups.
