## Supplement Figure S1 for "Extending influenza surveillance to detect non-influenza respiratory viruses of public health relevance: analysis of surveillance data, 2015-2019, Belgium"

**Supplement Figure S1:** Distribution of the positive samples for each respiratory virus by season and sampling week for ILI patients. Colour codes: black, tested by multiplex RT-qPCRs (Tested); light grey, influenza virus type A or B (Influenza); light pink, Respiratory Syncytial virus type A or B (RSV); fuchsia, human metapneumovirus (hMPV); green, parainfluenzavirus type 1, 2, 3 or 4 (PIV); orange, picornavirus of the *rhinovirus* and *enterovirus* genera or parechovirus (Pico); blue, coronavirus CoV-OC43, CoV-NL63 or CoV-229E (Corona); light brown, adenovirus (Adeno); purple, bocavirus (Boca). Plain bar, single detection; chequered pattern, in co-detection with at least one other virus. X-axis, weeks of active ILI surveillance: week 40 of year Y to week 20 of year Y+1.

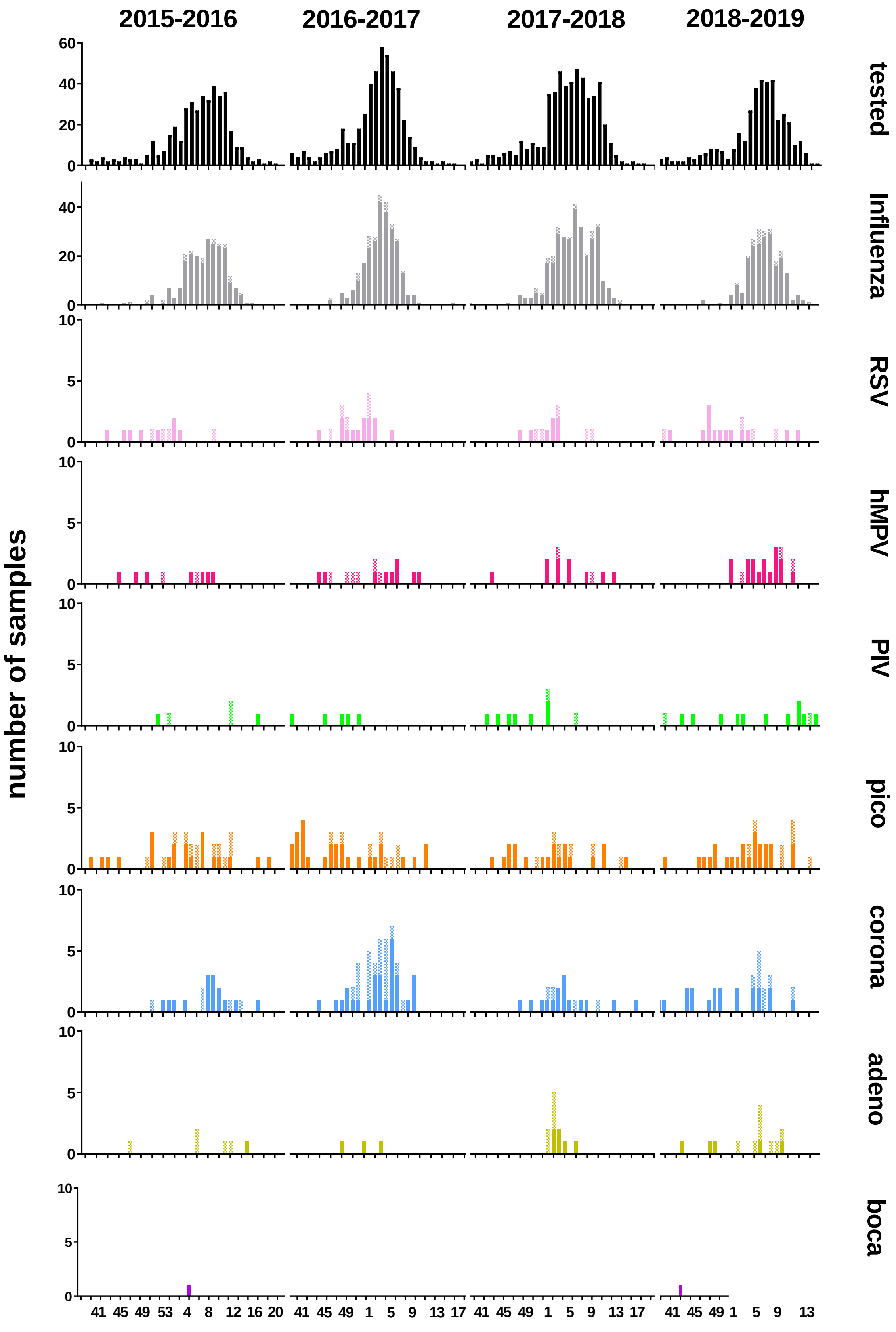

2015-2016
2016-2017
2017-2018
2018-2019
number of samples
tested
Influenza
RSV
hMPV
PIV
pico
corona
adeno
boca
60
40
20
0
10
5
41 45 49 53 4 8 12 16 20
41 45 49 1 5 9 13 17
41 45 49 1 5 9 13 17
41 45 49 1 5 9 13
